# Combined mitral and tricuspid TEER with a single TriClip steerable guide catheter: A single-center study

**DOI:** 10.1101/2025.01.28.25321297

**Authors:** Georgios E. Papadopoulos, Ilias Ninios, Sotirios Evangelou, Andreas Ioannides, Vlasis Ninios

## Abstract

**Background:** Combined M- and T-TEER typically involves two separate systems, complicating logistics and increasing procedural risks. This study aims to evaluate the safety and efficacy of combined mitral (M-TEER) and tricuspid (T-TEER) transcatheter edge-to-edge repair using a single TriClip® steerable guide catheter (SGC).

**Methods:** Patients with moderate-to-severe (3+) or severe (4+) degenerative (DMR) or functional (FMR) mitral regurgitation and massive/torrential or severe functional tricuspid regurgitation (TR), classified as New York Heart Association (NYHA) class III or IV, who underwent combined M- and T-TEER with the same TriClip SGC between January 2022 and December 2024, were included. The primary objectives included procedural outcomes, MR and TR severity reduction, and NYHA class improvement.

**Results:** Among 42 patients (64% female; median age: 77 years [IQR: 9]], the implantation success rate was 100%, with mean device and procedure times of 39.2 ± 6.9 and 71.2 ± 9.6 minutes, respectively. There were no in-hospital or 30-day major adverse events (MAEs), except for 2 patients (4.8%) with tricuspid single leaflet device attachment (SLDA), and 1 patient (2.4%) who underwent atrial septal defect (ASD) closure. Over a median follow-up period of 0.91 years, 3 (7.1%) patients were hospitalized for heart failure, with zero mortality. At 1-year follow-up, all patients achieved NYHA class ≤II, along with MR ≤2+ and 34 (81%) patients had only trivial/mild TR.

**Conclusions:** Combined M-TEER and T-TEER using the same TriClip SGC demonstrated exceptional safety and efficacy, along with significant functional and echocardiographic improvements.

## Introduction

Transcatheter edge-to-edge repair (TEER) has emerged as a promising therapeutic option, particularly for patients with mitral regurgitation (MR) who are at high surgical risk ^1–4^. Tricuspid regurgitation (TR) and MR often coexist ^1^, complicating the clinical picture and therapeutic strategy. Although current guidelines recommend surgical repair of TR in patients undergoing left heart cardiac surgery ^1^, robust data for high surgical risk patients is lacking.

Two retrospective studies compared the combined mitral (M-TEER) and tricuspid (T-TEER) transcatheter edge-to-edge repair versus T-TEER alone and both concluded in superiority of the combined method in terms of survival and functional improvements ^1–2^. However, this evidence comes from procedures with two dedicated systems, the steerable guide catheter (SGC) of Mitraclip® and TriClip® systems, respectively.

The two major differences between the Mitraclip and TriClip steerable guide catheters (SGCs) are the shorter curved tip of the TriClip SGC (1.5 cm vs. 2.5 cm) and, most importantly, the addition of a knob for septal-lateral steering. This additional knob facilitates optimal trajectory during T-TEER. Conversely, the longer curved tip of the Mitraclip SGC is considered advantageous for M-TEER on the left side. When performing a combined procedure using the two dedicated systems, the Mitraclip SGC must be retracted into the right atrium following Mitraclip implantation and then exchanged for the TriClip SGC to proceed with TriClip implantation. While this approach addresses anatomical specificity, it introduces logistical challenges. The exchange of two large-bore catheters may increase the risk of femoral venous trauma and bleeding complications.

We hypothesize that using the TriClip SGC for both valves is feasible, safe, and effective, enabling successful simultaneous M- and T-TEER. The shorter tip of the TriClip SGC, combined with the additional septal-lateral steering knob, may be well-suited for both valves and offer additional advantages. Notably, it may provide better maneuverability in challenging anatomies, such as patients with a small left atrium, where gaining procedural height is critical.

This is the first single center study to evaluate the procedural and mid-to-long term outcomes of combined M-TEER and T-TEER using the same SGC of the TriClip® system.

## Methods

This retrospective observational cohort study encompasses data extracted from medical records of all patients who underwent combined M-TEER and T-TEER at our structural heart disease expert center from January 2022 to January 2024. The study received approval from the Institutional Review Board of our center, in accordance with the Declaration of Helsinki.

### Inclusion Criteria

Patients diagnosed with moderate-to-severe (3+) or severe (4+) degenerative MR (DMR) or functional MR (FMR), combined with at least severe functional TR and classified as New York Heart Association (NYHA) class III or IV, who underwent combined M-TEER and T-TEER with the same SGC of the TriClip® system at our structural heart disease expert center between January 2022 and January 2024 were included. DMR was defined as MR resulting from a degenerative mitral valve disease, while FMR was characterized as MR stemming from compromised left ventricle or atria while the valve remained anatomically intact. The decision to perform the transcatheter procedure was made by the local Heart Team, guided by the 2021 European Society of Cardiology (ESC) / European Association for Cardio-Thoracic Surgery (EACTS) guidelines ^1^.

### Combined M-TEER and T-TEER procedure

The combined transcatheter edge-to-edge repair (TEER) procedure was performed under general anesthesia in the Hybrid Operating Room at the Interbalkan Medical Center, utilizing the MitraClip® and TriClip ® G4 system (Abbott Vascular, Santa Clara, CA, USA) by a single operator.

A fully percutaneous approach was employed for the procedure. After obtaining ultrasound-guided femoral venous access, a transseptal puncture was performed according to standard practice, targeting a mid-to-inferior and posterior location. The TriClip steerable guide catheter (SGC) was then advanced into the left atrium, and one or more MitraClip G4 devices were implanted under real-time fluoroscopic and 3D transoesophageal echocardiographic (TEE) guidance, following standard protocol. After successfully completing the M-TEER, the TriClip SGC was withdrawn into the right atrium. Subsequently, one or more TriClip G4 devices were implanted based on the patient’s anatomical requirements. Both the MitraClip and TriClip delivery systems were engaged with the SGC following the recommended alignment (“blue-to-blue” line), ensuring proper orientation.

A comprehensive echocardiographic assessment was performed to evaluate the final result. This included checking for residual MR and TR, assessing leaflet mobility, adequate leaflet grasping and confirming that no significant mitral stenosis had been induced. Hemodynamic measurements were repeated to assess the improvement in cardiac function post-procedure. Following the procedure, patients were transferred to the intensive care unit for close monitoring. Post-procedural TEE was typically performed within 24 hours to confirm the stability of the Mitra- and Tri-Clip and to reassess MR and TR severity.

### Definition of the variables

Baseline characteristics including age, sex, estimated glomerular filtration rate (eGFR), European System for Cardiac Operative Risk Evaluation II (EuroSCORE II), cardiopulmonary comorbidities (e.g., prior heart failure hospitalization, atrial fibrillation [AF], diabetes, chronic obstructive pulmonary disease [COPD]), cardiac resynchronization therapy (CRT), N-terminal pro-brain natriuretic peptide (NT-proBNP), and NYHA class were recorded. Transthoracic echocardiography (TTE) was conducted to assess left ventricular ejection fraction (LVEF), left ventricular end-diastolic diameter (LVDD), MR and TR severity. MR severity classification comprises of trivial (0+), mild (1+), moderate (2+), moderate-to-severe (3+), and severe (4+). TR severity classification comprises of trivial/mild, moderate, severe, massive/torrential. All TTEs were conducted by the same examiner. Right heart catheterization was performed before each procedure to measure mean right atrial pressure (mRAP), pulmonary capillary wedge pressure (PCWP), mean pulmonary artery pressure (mPAP), cardiac index (CI), pulmonary artery systolic pressure (PASP), and pulmonary vascular resistance (PVR). MR etiology was classified as DMR or FMR by the Heart Team.

### Study objectives

A TTE was performed at 30-day, and 1-year follow-up to assess MR and TR severity. NYHA class was also recorded at 30-day and 1-year follow-up. Survival and hospitalization data were obtained either through telephone contact or via the national electronic health record, when available. The main objectives of the study were to report procedural and mid-to-long term outcomes, i.e. to analyze the change in MR, TR severity and NYHA class throughout follow-up visits.

### Statistical analysis

Continuous variables following normal distribution were presented as mean and standard deviation (SD), while variables that were not distributed normally were presented as median and interquartile range (IQR). Normality of distribution was assessed by comparing mean and median values, graphical representation of the distribution of the variables and by using the Kolmogorov–Smirnov test. Qualitative variables were summarized using absolute and relative frequencies [n/N (%)]. Statistical comparisons of continuous variables that exhibited normal distribution were performed using the student t-test, while the Wilcoxon rank-sum test was employed for variables that did not follow a normal distribution. Categorical variables were compared with the χ2 test or the Fisher exact test if cell counts were small (≤5). The Kruskal-Wallis test was used for comparison of continuous variables between more than two independent samples. All statistical analyses were performed on RStudio version 2023.03.0+386.

## Results

### Baseline characteristics

In total, 42 patients (64% female, mean± SD age 77± 9 years) underwent combined M-TEER and T-TEER with the same TriClip SGC. Table 1 summarizes the baseline characteristics of the total cohort. Most of the patients were in advanced heart failure, symptomatic at rest with unfavorable hemodynamic profile.

**Table 1.** Patients’ baseline characteristics.

| Characteristic | N = 42<sup>1</sup> |
| --- | --- |
| Age, y | 77± 9 |
| Female sex | 27 / 42 (64) |
| NYHA class |  |
| III | 15 / 42 (35.7) |
| IV | 27 / 42 (64.3) |
| Euroscore II, % | 7± 2 |
| HF Hospitalization | 21 / 42 (50) |
| AF | 38 / 42 (90) |
| COPD | 19 / 42 (45) |
| Diabetes | 26 / 42 (62) |
| CRT | 4 / 42 (10) |
| eGFR, mL/min/1.73 m<sup>2</sup> | 45± 6 |
| NT-proBNP, pg/mL | 3,245 (1,828, 4,826) |
| *Echocardiography* |  |
| MR Etiology |  |
| DMR | 15 / 42 (35.7) |
| FMR | 27 / 42 (64.3) |
| MR severity |  |
| 3+ | 4 / 42 (10) |
| 4+ | 38 / 42 (90) |

Table 1. Patients' baseline characteristics
| Characteristic | N = 42<sup>1</sup> |
| --- | --- |
| TR severity |  |
| Severe | 32 / 42 (76) |
| Massive/Torrential | 10 / 42 (24) |
| LVEF, % | 43± 8.35 |
| LVDD, cm | 5.80± 1.02 |
| *Hemodynamics* |  |
| CI, mL/min/m<sup>2</sup> | 1.95± 0.6 |
| PASP, mmHg | 55± 10 |
| mPAP, mmHg | 33± 5 |
| PCWP, mmHg | 26± 6 |
| mRAP, mmHg | 15.0± 4 |
| PVR, WU | 3± 1.9 |

<sup>1</sup> Median (IQR); mean± SD; n / N (%)
Note: Continuous variables are presented as median value with interquartile range (IQR) or mean value± standard deviation (SD). Categorical variables are presented as n / N (%).
Abbreviations: NYHA: New York Heart Association, HF: Heart Failure, AF: Atrial Fibrillation, COPD: Chronic Obstructive Pulmonary Disease, CRT: Cardiac Resynchronization Therapy, eGFR: estimated Glomerular Filtration Rate, NT-proBNP: N terminal-pro brain natriuretic peptide, MR: Mitral Regurgitation, DMR: Degenerative Mitral Regurgitation, FMR: Functional Mitral Regurgitation, TR: Tricuspid Regurgitation, LVEF: Left Ventricular Ejection Fraction, LVDD: Left Ventricular End-Diastolic Diameter, CI: Cardiac Index, PASP: Pulmonary Artery Systolic Pressure, mPAP: mean Pulmonary Artery
Pressure, PCWP: Pulmonary Capillary Wedge Pressure, mRAP: mean Right Atrial Pressure, PVR: Pulmonary Vascular Resistance.

### Procedural Outcomes

Of the total 42 procedures, the implantation success rate was 100%, with median device and procedure times of 38.5± 7 minutes and 70± 10 minutes, respectively. The mean± SD length of hospital stay was 2.9± 0.3 days, and all patients (100%) were discharged home. There were no in-hospital or 30-day major adverse events (MAEs), i.e. death, stroke, myocardial infarction (MI), major bleeding, acute kidney injury (AKI), need for surgery, except for two patients (4.8%) with tricuspid single leaflet device attachment (SLDA), and one patient (2.4%) who underwent atrial septal defect (ASD) closure due to a right-to-left shunt (Table 2).

**Table 2.** Procedural Outcomes.

| **Procedural Outcomes** | **N = 42<sup>1</sup>** |
| --- | --- |
| Implantation rate | 42 / 42 (100) |
| Device time, mins | 38.5± 7 |
| Procedure time, mins | 70± 10 |
| Length of stay, days | 2.9± 0.3 |
| Discharged home | 42 / 42 (100) |
| Death | 0 / 42 (0) |
| Stroke | 0 / 42 (0) |
| MI | 0 / 42 (0) |
| Major Bleeding | 0 / 42 (0) |
| AKI | 0 / 42 (0) |
| SLDA (Tricuspid) | 0 / 42 (0) |
| Need for Surgery | 0 / 42 (0) |
| ASD closure | 1 / 42 (2.4) |

<sup>1</sup> Median (IQR); mean± SD; n / N (%)
Note: Continuous variables are presented as median value with mean value± standard deviation (SD). Categorical variables are presented as n / N (%).
Abbreviations: MI: Myocardial Infarction, AKI: Acute Kidney Injury, SLDA: Single Leaflet Device Attachment, ASD: Atrial Septal Defect

### Mid to long term outcomes

Over a median follow-up period of 0.91 years, 3 (7.1%) patients were hospitalized for heart failure, with zero deaths observed. At baseline, 27 (64.3%) patients were classified as NYHA class IV, with 11 (40.7%) of them being fully asymptomatic in 30 days (Figure 1). At 1-year follow-up, all patients achieved NYHA class I or II.

**Figure 1.**
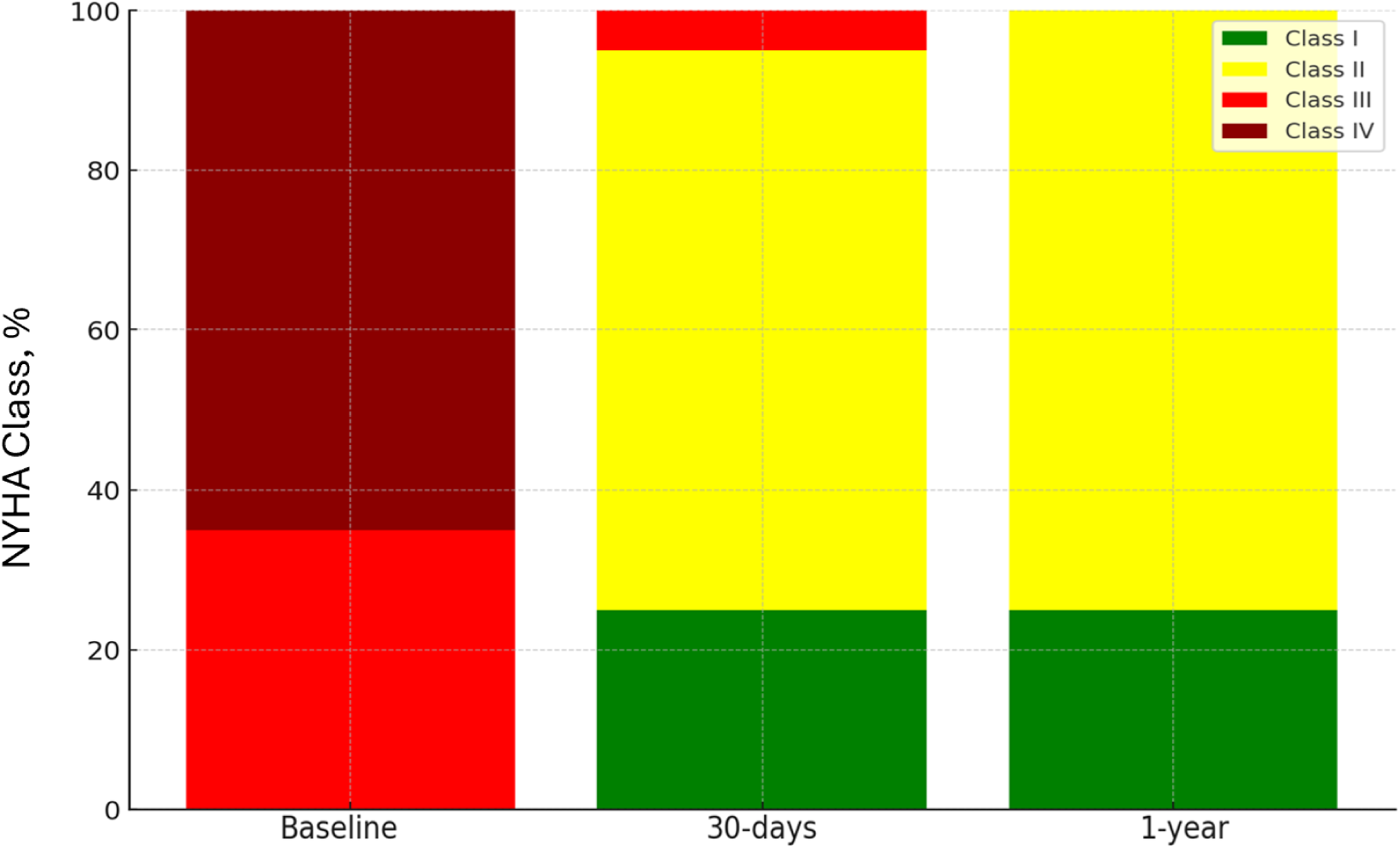
NYHA Class distribution over time. At 1-year follow-up, all patients achieved NYHA class I or II. Abbreviations: NYHA: New York Heart Association

Thirty-eight (90%) patients showed severe (4+) MR at baseline TTE. At 30-days and 1-year follow-up, MR ≤2+ was achieved in all patients, with 27 (71.1%) severe MR patients showing only mild MR after 1 year (Figure 2).

**Figure 2.**
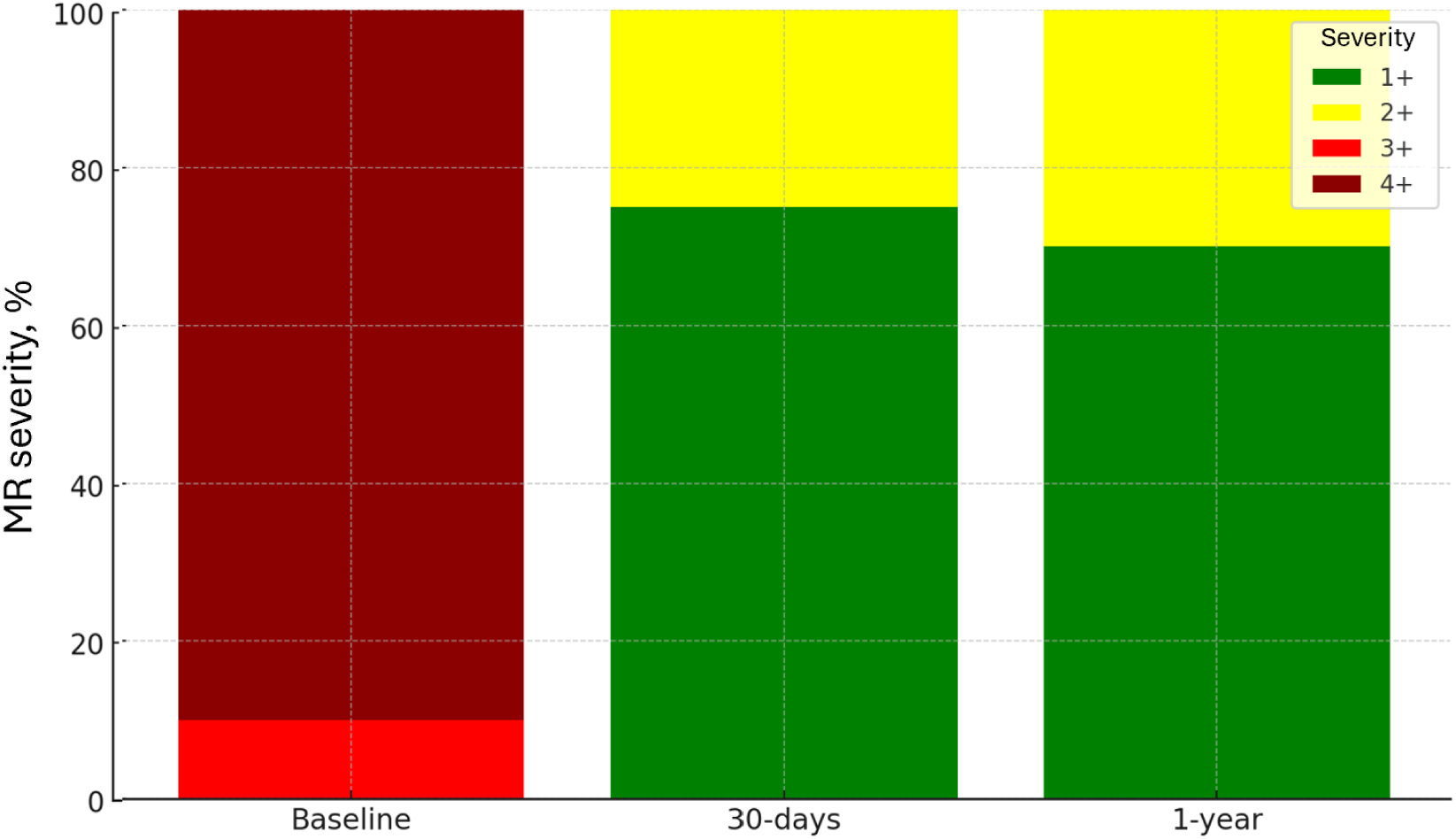
MR severity distribution over time. At 30-days and 1-year follow-up, MR ≤2+ was achieved in all patients. 1+: Mild MR, 2+: Moderate MR, 3+: Moderate-to-severe MR, 4+: Severe MR. Abbreviations: MR: Mitral Regurgitation

Similarly, out of 42 massive/torrential or severe TR patients at baseline, in 30-days and 1-year follow-up, 34 (81%) patients had only trivial/mild TR and 5 (12%) had moderate TR (Figure 3).

**Figure 3.**
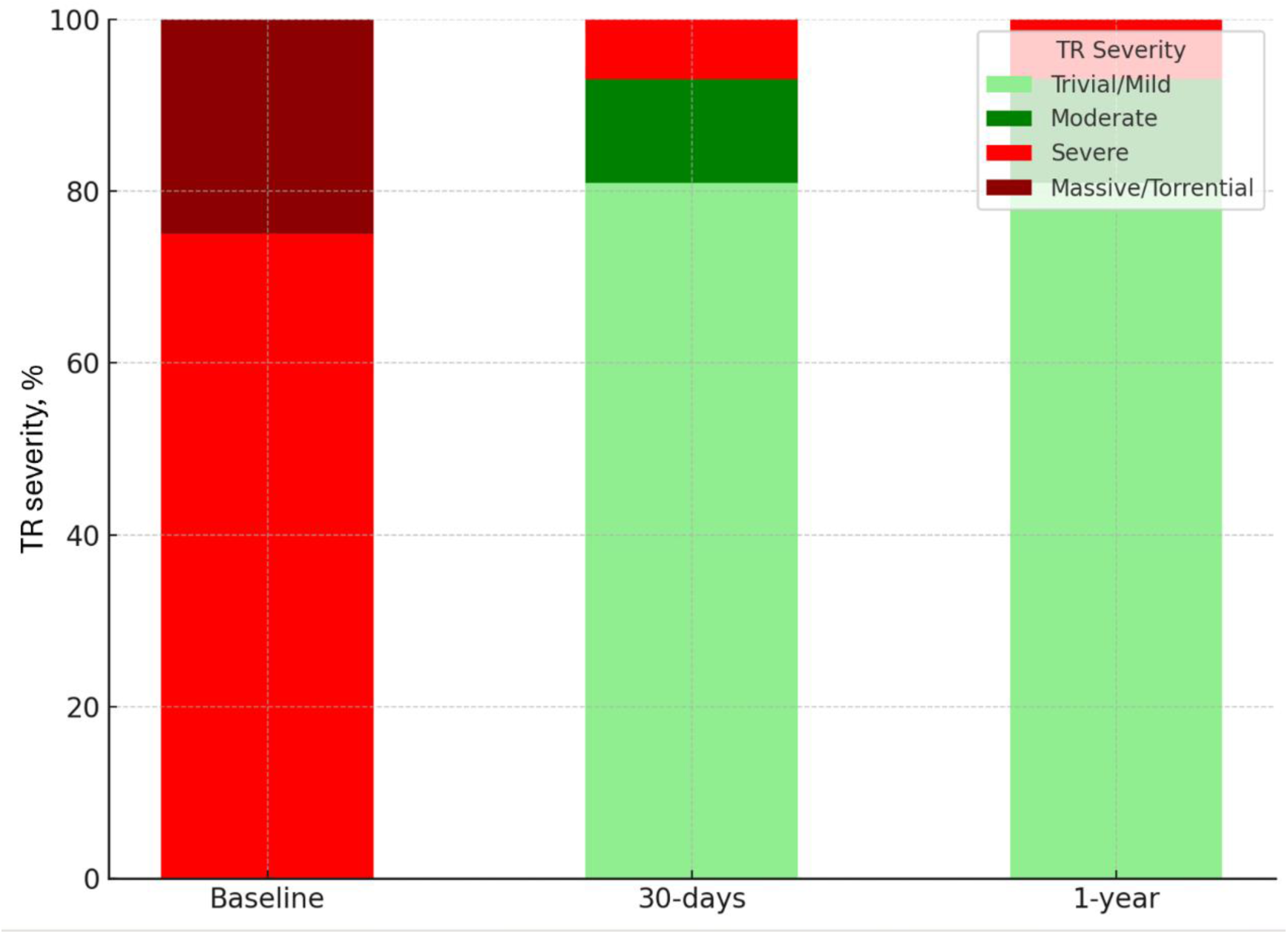
TR severity distribution over time. In 30-days and 1-year follow-up, 39 (93%) patients had moderate or less TR. Abbreviations: TR: Tricuspid Regurgitation

## Discussion

This single-center study provides an analysis of the procedural and mid-to-long-term outcomes of combined M-TEER and T-TEER using a single TriClip SGC system, demonstrating excellent safety and efficacy, along with significant improvements in both functional and echocardiographic parameters. Additionally, heart failure hospitalization rates were low, with no mortality observed during the follow-up period.

To our knowledge, this is the first comprehensive study to report the use of a single SGC system for both M-TEER and T-TEER. A prior first-in-human series described seven cases using the TriClip SGC for combined M- and T-TEER, demonstrating the feasibility of the approach ^4^. Our findings extend this preliminary evidence by providing detailed short- and mid-term outcomes, including a larger patient cohort and a thorough evaluation of clinical improvements over a one-year period.

The results align with existing data, indicating that combined mitral and tricuspid repair offers superior functional and survival outcomes compared to isolated valve interventions ^7–8^. In our cohort, there was a significant reduction in both MR and TR severity. Specifically, MR decreased from severe to moderate or mild in all patients at 30 days, and this improvement was sustained at the 12-month follow-up. Likewise, TR was reduced from massive/torrential or severe to moderate or less in 93% of the patients. These findings, consistent with previous studies ^7^, demonstrate that the combined approach provides durable reductions in both MR and TR, effectively alleviating the burden of valvular dysfunction.

In terms of functional improvement, all patients experienced substantial advancements in NYHA functional class, with all moving to class I or II from a baseline of class III or IV, likely attributable to the simultaneous resolution of both valvular insufficiencies. The functional improvement seen with the single-system approach exceeded that reported with the use of two dedicated systems, where only 67% of patients reached NYHA class ≤II at one year ^8^. These clinical benefits were further underscored by zero mortality and a reduction in heart failure hospitalizations, with only 7.1% of patients requiring readmission within the first year. Notably, previous studies using the two-system approach reported a 1-year mortality rate of 16% ^8^ and 18-month heart failure hospitalization and mortality rates of 5.5% and 33.3%, respectively ^7^.

A key advantage of the single SGC approach was the reduction in procedural complexity. The ability to perform both M-TEER and T-TEER without the need for catheter exchanges or additional venous punctures streamlined the procedure. The average procedural time was approximately half of the range reported for staged mitral and tricuspid interventions. Importantly, none of the patients in our cohort experienced procedural complications, such as access-site bleeding, stroke, or acute kidney injury-complications often seen in this high-risk population. This underscores the safety of the single-system approach and its potential to minimize procedural risks, even in patients with multiple comorbidities. Technically, using the TriClip SGC on the left side not only avoided additional technical challenges but was potentially advantageous in patients with small left atria and a low fossa ovalis position, where achieving adequate height has historically been challenging. In a subset of patients, the use of the septal flexion (“S”) knob provided additional height without compromising the trajectory toward the mitral valve plane.

Despite these promising results, this study has limitations. The single-center, retrospective design and relatively small sample size may limit the generalizability of our findings. However, it is important to note that our center functions as a referral hub for structural heart disease patients across a broader region in northern Greece, mitigating potential selection bias. Furthermore, while mid-term outcomes are favorable, longer-term follow-up is necessary to evaluate the durability of the valve repairs and their impact on long-term survival and quality of life. Larger, multicenter, randomized trials are needed to validate our findings and to establish optimal patient selection criteria for the single-system approach.

In conclusion, our study provides a comprehensive evaluation of the safety and efficacy of a single SGC system for combined M-TEER and T-TEER. This approach simplifies the procedure, reduces procedural complexity, and results in significant and sustained improvements in valve function and clinical outcomes, with minimal adverse events. In conjunction with the first-in-human series, our findings emphasize the potential of this technique for managing high-risk patients with severe dual-valve disease. Future studies should aim to expand the applicability of this approach and confirm its long-term benefits in a larger and more diverse patient population.

## Data Availability

The data that support the findings of this study are available from the corresponding author upon reasonable request.

## Acknowledgements

The authors thank the staff of the Interbalkan Medical Center for their support and collaboration during this case.

## Source of Funding

No external funding was received.

## Disclosures

The authors have no relevant financial relationships or conflicts of interest to disclose.

## Non-standard Abbreviations and Acronyms

MR: Mitral Regurgitation
TR: Tricuspid Regurgitation
SGC: Steerable Guide Catheter
TEER: Transcatheter Edge-to-Edge Repair
NYHA: New York Heart Association
FMR: Functional Mitral Regurgitation
DMR: Degenerative Mitral Regurgitation
eGFR: Estimated Glomerular Filtration Rate
NT-proBNP: N-terminal Pro-brain Natriuretic Peptide
EuroSCORE II: European System for Cardiac Operative Risk Evaluation II
AF: Atrial Fibrillation
COPD: Chronic Obstructive Pulmonary Disease
CRT: Cardiac Resynchronization Therapy,
TTE: Transthoracic Echocardiography
TEE: Transesophageal Echocardiography
LVEF: Left Ventricular Ejection Fraction
LVDD: Left Ventricular end-Diastolic Diameter
CI: Cardiac Index
PASP: Pulmonary Artery Systolic Pressure
mPAP: mean Pulmonary Artery Pressure
PCWP: Pulmonary Capillary Wedge Pressure
mRAP: mean Right Atrial Pressure
PVR: Pulmonary Vascular Resistance
MI: Myocardial Infarction
AKI: Acute Kidney Injury
SLDA: Single Leaflet Device Attachment
ASD: Atrial Septal Defect
MAE: Major Adverse Event

## Notes

### Competing Interest Statement

The authors have declared no competing interest.

### Author Declarations

Institutional Review Board of European Interbalkan Medical Center Approval Number: 2335 Date of Approval: September 09, 2024

